# Temporal Variations in Ischemic and Bleeding Event Risks after Acute Coronary Syndrome during Dual Antiplatelet Therapy

**DOI:** 10.1101/2023.04.25.23289123

**Authors:** Toshiharu Fujii, Satoshi Kasai, Yota Kawamura, Fuminobu Yoshimachi, Yuji Ikari

**Author notes:** Address for correspondence Toshiharu Fujii M.D., Ph.D., Address: Department of Cardiovascular Medicine, Tokai University School of Medicine 143 Shimokasuya, Isehara, 259-1193, Japan.

## Abstract

**Background:** This study estimates the temporal risk variations of ischemic and bleeding events during dual antiplatelet therapy (DAPT) and suggests the optimal period for DAPT after acute coronary syndrome (ACS) categorized by the Academic Research Consortium for High Bleeding Risk (ARC-HBR) criteria.

**Methods:** A total of 1,264 ACS patients receiving either clopidogrel or prasugrel with aspirin were classified by ARC-HBR criteria into HBR (n = 574) and non-HBR groups (n = 690). This study was designed as a multicenter observation to evaluate the primary endpoints of ischemic, including cardiogenic death, myocardial infarction, or ischemic stroke, and bleeding events, defined as Bleeding Academic Research Consortium type 3 or 5. The temporal risk variations were estimated using the Cox hazard and Royston-Parmar models.

**Results:** Ischemic and bleeding events were observed in 9.4% and 7.4% of patients, respectively, during an average observation period of 313 days. The HBR group had a higher incidence of both events than the non-HBR group (15.3% vs. 4.5%, P < 0.01 for ischemic; 11.9% vs. 3.8%, P < 0.01 for bleeding). The estimated risk curves revealed a peak and steep decline in the first few days, followed by a constant decline. The peak of risk was higher for bleeding than for ischemic events, but this relationship reversed early, with ischemic events displaying a higher risk in both the HBR and non-HBR groups until at least 60 days.

**Conclusion:** A 60-day period of DAPT is appropriate to balance the risks of adverse events after ACS, regardless of ARC-HBR criteria.

## 1. Introduction

Antiplatelet agents play an important role in reducing ischemic events in patients after ischemic heart disease. Prolonged dual antiplatelet therapy (DAPT) increases the risk of adverse bleeding events, particularly in cases of acute coronary syndrome (ACS). Short-term DAPT is becoming a more popular approach, and the current focus is on minimizing the duration of antiplatelet therapy while reducing the risk of both bleeding and ischemic events.

The Academic Research Consortium for High Bleeding Risk (ARC-HBR) has developed standardized criteria to identify patients at high risk of bleeding and to allow for selection of appropriate antiplatelet regimens.^1,2^ Several clinical guidelines recommend shorter periods of DAPT for patients who meet the ARC-HBR criteria compared to those with a lower risk. ^3,4^

Although numerous prospective and histological studies had examined the optimal period of DAPT, the available clinical evidence on short-term DAPT is limited, consisting mainly of fragmented comparisons or isolated case reports. Furthermore, the currently available clinical evidence is insufficient to enable determination of the optimal period of short-term DAPT based on ARC-HBR criteria.

To determine the optimal period of DAPT for ACS patients who are categorized by ARC-HBR criteria, the present study estimates the temporal variations in the risk of adverse ischemic and bleeding events during DAPT.

## 2. Methods

### 2.1. Study design and population

The present study was designed as a multicenter observational study and conducted on 1,551 patients with ACS who were admitted to either Tokai University School of Medicine or Tokai University Hachioji Hospital between May 2014 and May 2022. Patients who received DAPT, either clopidogrel (75 mg daily maintenance dose) or prasugrel (3.75 mg daily maintenance dose) in addition to aspirin (100 mg daily maintenance dose) after admission were surveyed using existing medical records. Patients who received antiplatelet or anticoagulation therapy other than aspirin and clopidogrel or prasugrel were excluded from the study cohort. A total of 1,264 patients meeting these requirements were divided into two groups based on whether they fulfilled the ARC-HBR criteria at hospital arrival: HBR (n = 574) and non-HBR (n = 690).

The primary outcomes of the study were occurrences of ischemic and bleeding events during the observation period. To define the optimal period for DAPT after ACS, the temporal variations in risk of ischemic and bleeding events were analyzed using the Cox and Royston-Parmar models in both the HBR and non-HBR groups. The Institutional Review Board for Clinical Research of Tokai University Hospital approved this study (22R496).

### 2.2. Outcomes and definitions

ACS was defined according to the Fourth Universal Definition of Myocardial Infarction. ^5^ ARC-HBR scoring was used to assess each individual ‘s bleeding risk upon hospital arrival. ^1^ Ischemic events consisted of cardiogenic death, myocardial infarction, and ischemic stroke. Bleeding events were defined as Bleeding Academic Research Consortium (BARC) types 3 or 5.^6^ An ischemic stroke was defined as the sudden onset of new neurologic deficits attributed to the obstruction of cerebral blood flow with no apparent non-vascular cause, as confirmed by imaging.

The observation period was defined as the time between the start of dual antiplatelet therapy and any change or discontinuation of the therapy regimen for the patient.

### 2.3. Statistical analysis

Numerical data with a normal distribution were presented as mean ± standard deviation, and a Student ‘s t-test was used to assess any significant differences between the two groups. A Fisher ‘s test was used to determine statistically significant differences between categorical variables.

Time to event was analyzed using the Cox proportional-hazards model. The proportional-hazards assumption was tested using Schoenfeld residuals after fitting a model with the Cox proportional hazards regression models. The following parameters were used to adjust the confounders: age, sex, hypertension, dyslipidemia, diabetes mellitus, hemoglobin, platelet count, eGFR, hemodialysis, diagnosis, shock vital at the time of arrival to hospital, OHCA, and usage of mechanical support devices. The hazard function was estimated using a kernel smooth of the estimated hazard contributions.

To calculate the hazard function of the standardized survival curve for ischemic and bleeding events, the Royston-Parmar model, a flexible parametric model fitted on the hazard scale with restricted cubic splines, was used. This model estimated the average HR, as well as the temporal pattern of HR, together with their 95% CIs. ^7,8^ The following parameters were used to establish the parametric model: age, sex, hypertension, dyslipidemia, diabetes mellitus, hemoglobin, platelet count, eGFR, hemodialysis, diagnosis, shock vital at the time of arrival to hospital, OHCA, and usage of mechanical support devices. The baseline hazard was modeled using a restricted cubic spline, and the units in this analysis were cases per 1000 person-years.

P values less than 0.05 were considered statistically significant, and all statistical calculations were performed using STATA statistical software, version 17.0 (Stata Corporation, College Station, TX, USA).

## 3. Results

### 3.1. Baseline characteristics and outcomes

To determine the optimal period of DAPT for ACS patients who are categorized as HBR by ARC-HBR criteria, 1264 patients were classified as HBR (n = 574) or non-HBR (n = 690) and their temporal variations in the risk of adverse ischemic and bleeding events during DAPT were estimated.

Table 1 summarizes the baseline characteristics of the study population. During the average 313.1 ± 322.4 days of the observation period (265.9 ± 318.0 days in HBR and 352.3 ± 321.1 in the non-HBR groups), ischemic and bleeding events were observed in 9.4% (15.3% in HBR vs 4.5% in non-HBR, P < 0.01; HR 4.6, 95% CI 2.8-7.4, P < 0.01) and 7.4% (11.9% in HBR vs 3.8% in non-HBR, P < 0.01; HR 3.5, 95% CI 2.1-5.7, P < 0.01) of patients, respectively.

**Table 1.** Baseline characteristics

|  | Overall<br>n = 1,264 | HBR<br>n = 574 | Non-HBR<br>n = 690 | P value |
| --- | --- | --- | --- | --- |
| Age, years | 68.9 ± 12.1 | 75.7 ± 10.0 | 63.2 ± 10.6 | <0.01 |
| Male, n | 999 (79.0%) | 425 (74.0%) | 574 (83.2%) | <0.01 |
| Hypertension, n | 943 (74.6%) | 451 (78.6%) | 492 (71.3%) | <0.01 |
| Dyslipidemia, n | 909 (71.9%) | 378 (65.6%) | 531 (77.0%) | <0.01 |
| Diabetes mellitus, n | 517 (40.9%) | 253 (44.1%) | 264 (38.3%) | 0.04 |
| Hemoglobin, g/dl | 13.7 ± 2.2 | 12.2 ± 2.1 | 15.0 ± 1.4 | <0.01 |
| Platelets, ×10<sup>4</sup>/μl | 22.3 ± 11.0 | 20.7 ± 11.6 | 23.6 ± 10.3 | <0.01 |
| Serum creatinine, mg/dl | 1.6 ± 2.1 | 2.5 ± 2.9 | 0.9 ± 0.2 | <0.01 |
| Estimated GFR, ml/min/1.73 m<sup>2</sup> | 55.9 ± 24.7 | 41.3 ± 25.4 | 68.1 ± 15.8 | <0.01 |
| Hemodialysis, n | 113 (8.9%) | 113 (19.7%) | 0 | <0.01 |
| Diagnosis of STEMI, n | 685 (54.2%) | 257 (44.8%) | 428 (62.0%) | <0.01 |
| Combined use with aspirin |  |  |  |  |
| Prasugrel, n | 734 (58.1%) | 265 (46.2%) | 469 (68.0%) | <0.01 |
| Clopidogrel, n | 530 (41.9%) | 309 (53.8%) | 221 (32.0%) |  |
| Observation period, days | 313.1 ± 322.4 | 265.9 ± 318.0 | 352.3 ± 321.1 | <0.01 |

Data are presented as the mean ± standard deviation, unless otherwise indicated.
GFR, glomerular filtration rate; STEMI, ST-elevation myocardial infarction; OHCA, out-of-hospital cardiac arrest

### 3.2. Cox proportional hazards regression models

Cox proportional hazards regression models were used to demonstrate the time-dependent changes in the risk of ischemic and bleeding events between the HBR and non-HBR groups (Figure 1). The HBR group had a higher risk of both events, particularly in the first few weeks. However, the risk of both events remained constant over time, and no major difference was observed between the two groups.

**Figure 1.**
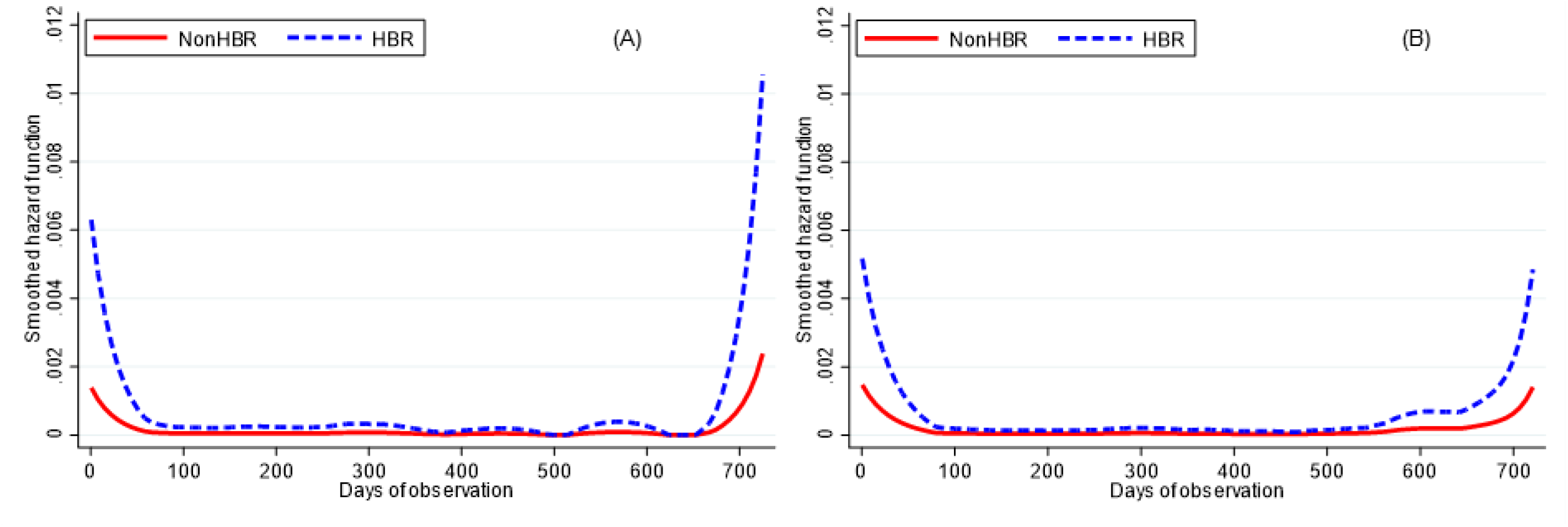
Hazard functions of ischemic and bleeding events

### 3.3. Flexible parametric models

Flexible parametric models were also used to demonstrate the temporal variations in the risk of ischemic and bleeding events (Figure 2). Occurrences of both events showed early initial peaks, followed by steep, then gradual decreases.

**Figure 2.**
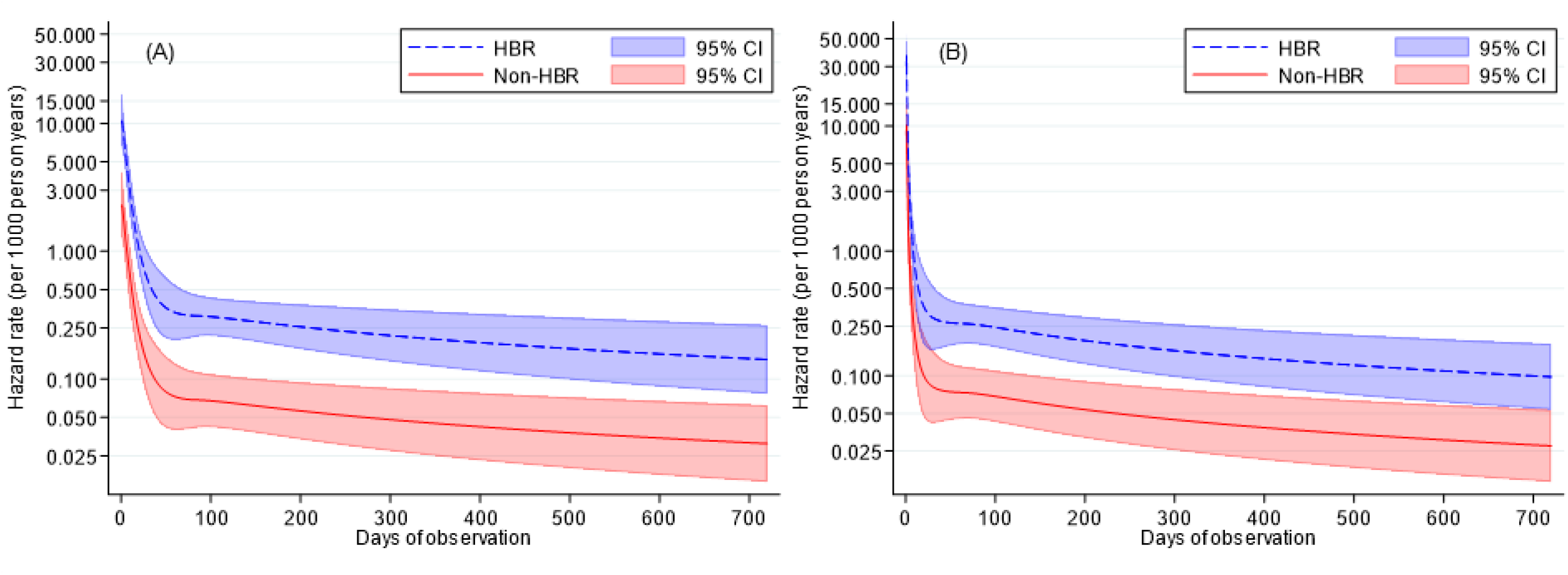
Temporal variations in risks of ischemic and bleeding events The temporal variations in the hazard rates with 95% confidence intervals for ischemic (A) and bleeding events (B) are presented separately for the HBR and non-HBR groups.

In Figure 3, the first 90 days of Figure 2 are focused on, and the risk of ischemic and bleeding events in the HBR and non-HBR groups was compared for this specific time window. The peaks of bleeding event risk were higher than those of ischemic event risk in both groups. The risk of ischemic events exceeded that of bleeding events several days after the peak due to a steeper decline in bleeding events than in ischemic events. In the non-HBR group, the risks of ischemic and bleeding events became equivalent after about 60 days and remained parallel thereafter. In the HBR group, although the difference between the risks of ischemic and bleeding events decreased until around day 60, the risk of ischemic events remained slightly higher even after this time.

**Figure 3.**
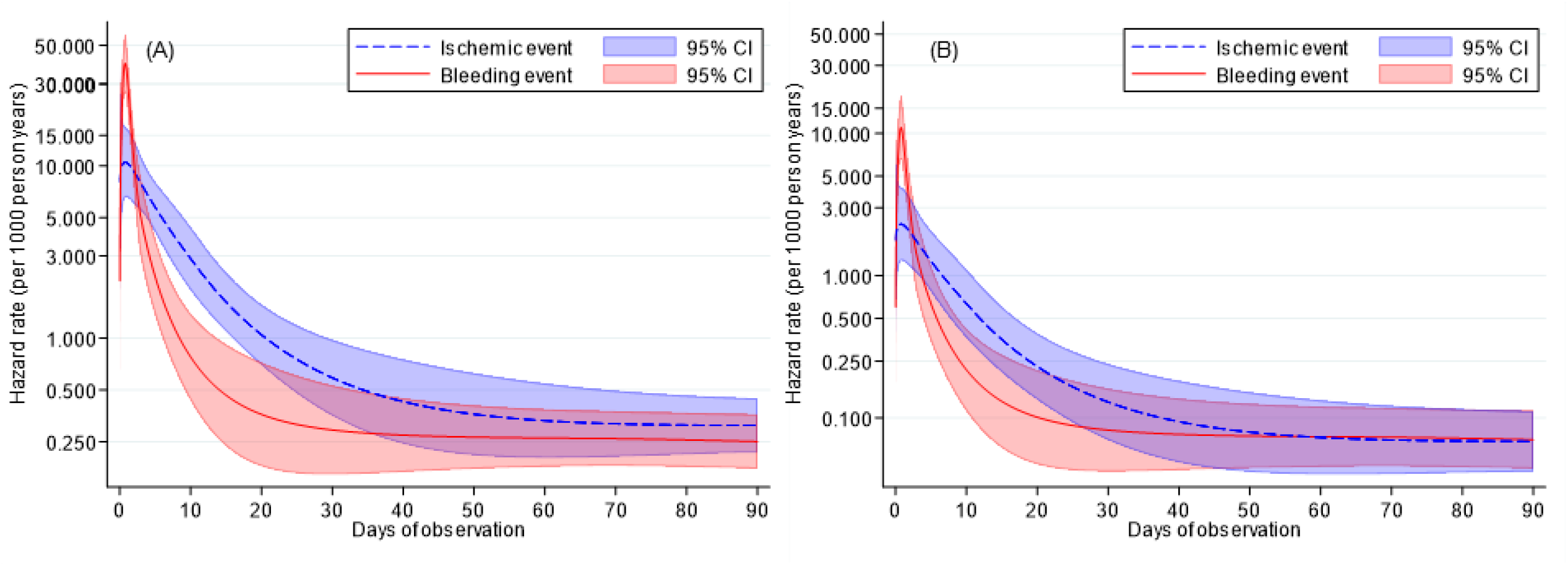
Temporal variations in risks focused on the first 90 days An enhanced look at the first 90 days of Figure 2 and comparing the risk of ischemic and bleeding events, which are shown separately as the HBR (A) and the non-HBR groups (B). Individual hazard rates per 1000 person years at 1, 5, 10, 20, 30, 40, 50, 60, 70, 80, and 90 days of observation are as follows: A) HBR group Ischemic event; 10.42, 5.77, 2.90, 1.05, 0.59, 0.43, 0.36, 0.33, 0.32, 0.31, and 0.31. Bleeding event; 35.21, 2.28, 0.78, 0.36, 0.29, 0.27, 0.27, 0.26, 0.26, 0.26, and 0.25. B) Non-HBR group Ischemic event; 2.29, 1.27, 0.64, 0.23, 0.13, 0.09, 0.08, 0.07, 0.07, 0.69, and 0.68. Bleeding event; 9.85, 0.64, 0.22, 0.10, 0.08, 0.08, 0.07, 0.07, 0.07, 0.07, and 0.07.

## 4. Discussion

The present study estimated the temporal variations in the risk of adverse ischemic and bleeding events during DAPT to determine the optimal period of DAPT for patients after ACS who are categorized as high-risk by ARC-HBR criteria.

In both the HBR and non-HBR groups, occurrences of both adverse events had early peaks followed by steep decreases, then remained at relatively low levels. The bleeding risk peaked in the first 1-2 days and was higher than the peak of ischemic risk. The HBR group had higher risks of both events than the non-HBR group throughout the observation period. The peak of ischemic risk in the HBR group was almost equivalent to the peak of bleeding risk in the non-HBR group. The temporal relationship between bleeding and ischemic risks reversed soon after the initial peaks. The decrease in risk after the first peak was steeper for bleeding events than for ischemic events, and this trend persisted for about 20 days for bleeding events and 60 days for ischemic events. After that, risk levels became constant, and the order of ischemic and bleeding risks reversed at 60 days in the non-HBR group. However, this inversion was not observed in the HBR group, where the parallel relationship persisted, with slightly higher ischemic than bleeding risk, even after 60 days. Based on these findings, we recommend a 60-day period of DAPT for patients not fulfilling the HBR criteria, considering the balance between efficacy and safety. Even for the HBR group, a 60-day DAPT is also deemed reasonable after the interpretation of these results.

The present study is novel in that it visualized the temporal variations in adverse events categorized by ARC-HBR using these estimated curves. This allows for a more informed recommendation by balancing the risk of both ischemic and bleeding events after ACS. Another noteworthy finding was that the HBR group had a higher risk of both bleeding and ischemic event than the non-HBR group throughout the observation period.

The latest guidelines from the European Society of Cardiology (ESC) recommend different treatment approaches for stable coronary artery disease and ACS. ^9^ For ACS, the recommended period of DAPT is 12 months, reduced to 6 months for patients with a high bleeding risk as assessed by the HAS-BLED score. For patients without ST-segment elevation, the ESC guidelines recommend 12 months of DAPT, but 3 months or 1 month for subgroups with high or very high bleeding risks, respectively. ^4^ The Japanese Circulation Society previously used the ESC regimen for patients with coronary artery disease but revised it in 2020 to assess bleeding risk using the modified ARC-HBR criteria. ^3^ The guidelines recommend 1 to 3 months of DAPT in patients with ARC-HBR, despite limited clinical evidence supporting short-term DAPT.

The SMART-CHOICE Randomized Clinical Trial compared major adverse events between 12 months and 3 months of DAPT followed by clopidogrel monotherapy and showed that the reduced 3-month regimen of DAPT is not statistically inferior in terms of major adverse cardiac and cerebrovascular events but is superior in terms of preventing adverse bleeding events. ^10^ Furthermore, the STPDAPT-2 trial also suggested that 1 month of DAPT had a better outcome than 12 months in terms of a composite of adverse ischemic and bleeding events. ^11^ Although these trials support short-term DAPT recommended in national guidelines, the results are limited and fragmented in these comparative studies contrasting 12 months.

Several studies have examined the percentage of neointimal coverage after implantation of current drug-eluting stent through pathological or imaging modalities. These studies have reported that neointimal coverage is approximately 85% after one month and around 90% after three months. ^12 13^ Some reports have suggested that short-term DAPT is safe in terms of the risk of stent thrombosis based on pathological findings. However, the problem with these prospective comparative or pathological studies is that they involve fragmented data or only a few cases, making it difficult to draw conclusions about the optimal period of DAPT. It is unclear whether these findings can be generalized to all patients after ACS.

The present study found that whether or not patients met the ARC-HBR criteria had a significant impact on their baseline characteristics, as shown in Table 1. ARC-HBR is well-known for its ability to predict bleeding events, even in cases of ACS. ^14^ While this was also demonstrated in the present study, the HBR group had a higher rate of not only bleeding events, but also ischemic events. Therefore, the HBR group had a higher overall risk of both ischemic and bleeding risk compared to the non-HBR group. ^15^ The present study also found that bleeding risk remained higher in the HBR group throughout the observation period. Furthermore, bleeding was concentrated in the first several days, regardless of whether patients met the ARC-HBR criteria or not. Given that the period of peak of bleeding risk is the only time when bleeding risk is higher than the risk of ischemia throughout the observation period, close attention should be paid to bleeding events during the first several days.

Conversely, the period after the peak of bleeding should be a time for attention to ischemic events. From the perspective of short-term DAPT, the present study recommends that non-HBR group should continue DAPT for at least 60 days, since their ischemic risk is higher than their bleeding risk until the 60 days mark—except for during the high early peak of bleeding risk—and almost equal to these risks after 60 days. In the HBR group, there is no period which bleeding risk exceeds ischemic risk throughout the observation period after 60 days. Therefore, it is reasonable to discontinue DAPT at 60 days when the risk of ischemia decreases and stabilizes.

The study had some limitations. The sample size might not have been sufficient to evaluate major adverse events. Also, patients with early censored observations could include a considerable number of low-risk patients, which might have resulted in higher event rates present in these data than in a typical ACS population.

In conclusion, the present study estimated the temporal variations in the risk of adverse ischemic and bleeding events during DAPT and recommends discontinuing DAPT at 60 days for patients after ACS who satisfy ARC-HBR criteria as well as those who do not.

## Data Availability

Data Availability; The deidentified participant data will not be shared.

## Acknowledgments

No Acknowledgement.

